# Human Tear Film Protein Sampling Using Soft Contact Lenses

**DOI:** 10.1101/2023.02.09.23285551

**Authors:** Robert K. Roden, Nathan Zuniga, Joshua C. Wright, David H. Parkinson, Leena M. Patil, Fangfang Jiang, Rebecca S. Burlett, Joshua J. Rogers, Jarett T. Pittman, Connor O. Roper, Kenneth L. Virgin, P. Christine Ackroyd, John C. Price, Kenneth A. Christensen

## Abstract

Human tear protein biomarkers are useful for detecting both ocular and systemic diseases. Unfortunately, existing tear film sampling methods (Schirmer strips [SS] and microcapillary tubes [MCT]) have significant drawbacks. Here we present an alternative tear protein sampling method using soft contact lenses (SCLs). First, we optimized SCL protein sampling *in vitro*, then performed *in vivo* studies in 6 human subjects. Using Etafilcon A SCLs and 4M guanidine-HCl for protein removal, we sampled an average of 60±31 μg of protein per eye. We also performed objective and subjective assessments of all sampling methods. Proteomic analysis by mass spectrometry (MS) revealed the majority of proteins were sampled by all methods. However, smaller subsets of unique and shared proteins were identified, particularly for SS and MCT. Additionally, SCLs had reduced levels of reflex tear protein zymogen granule protein 16 homolog B (ZG16B) versus SS and MCT. These experiments demonstrate SCLs as an accessible tear sampling method which may minimize reflex tearing and potentially aid biomarker discovery.

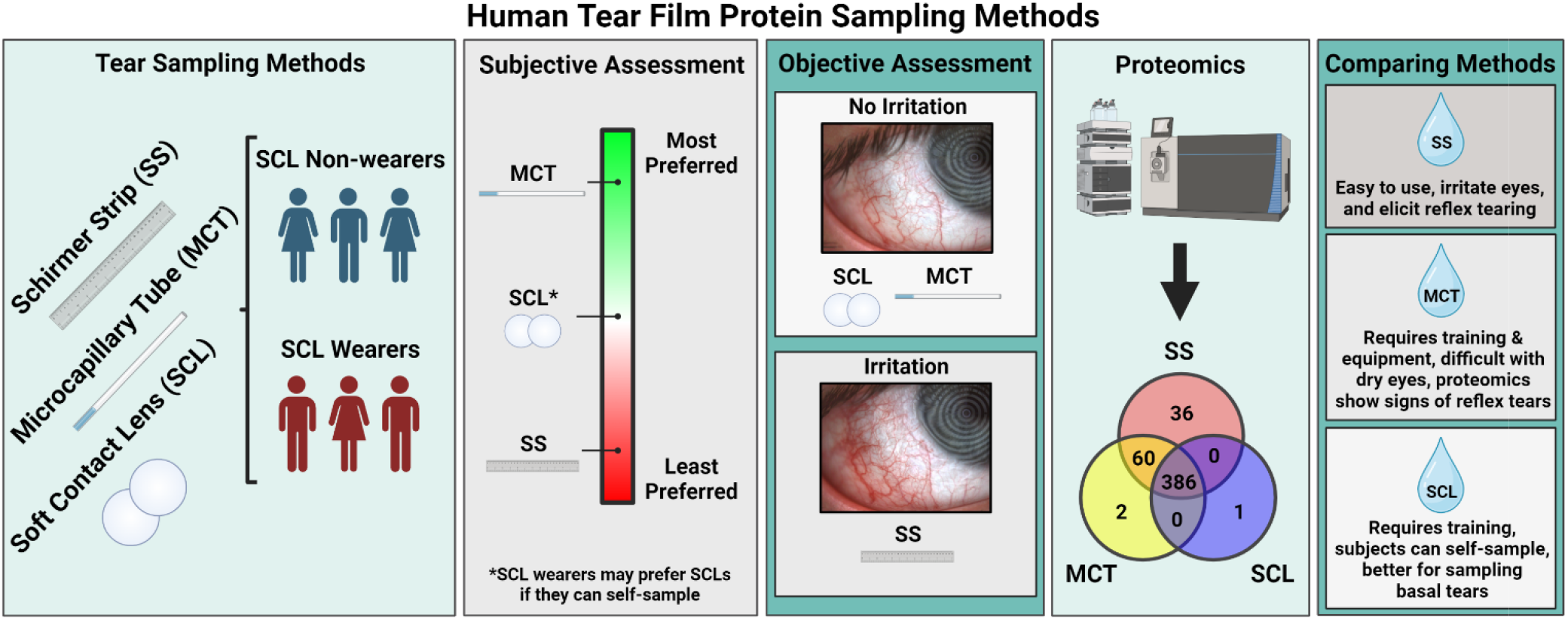

## Introduction

Human tear film is an attractive biospecimen given its accessibility and potential for use in diagnostic screenings [1]. In recent years, human tear protein biomarkers have been identified for various diseases, including glaucoma, multiple sclerosis, Parkinson’s disease, Alzheimer’s disease, and various forms of cancer [2,3]. Mass spectrometric identification of relevant disease biomarkers could lead to clinical diagnostics directly from tears [3,4]. However, a considerable obstacle to tear analysis is sampling. The most common tear sampling methods are cellulose filter Schirmer strips (SS) and microcapillary tubes (MCT) [5]. Each method has considerable drawbacks, including pain, irritation, damage to the conjunctival epithelium, risk of corneal injury, difficulty in capturing tears, and/or low sample volume [6,7]. Furthermore, the choice of tear sampling method impacts downstream analysis because of proteomic differences between tear sampling methods [8,9]. Thus, improved tear sampling methods are needed.

Soft contact lenses (SCLs) are FDA-approved hydrogels commonly used for vision correction. During contact lens wear, SCLs capture and concentrate proteins present in tear film by adsorption and absorption [10,11]. We hypothesized that if such proteins could be sampled and analyzed, SCLs would provide an alternative tear sampling method suitable for proteomic biomarker discovery and potentially diagnostics. As SCLs are designed for interaction with the ocular surface and optimized for comfort, we further hypothesized that SCLs would have advantages over current tear sampling methods.

Here we present an objective and subjective analysis of SCLs as a tear protein sampling method and compare these results to SSs and MCTs. These data demonstrate that SCLs yield comparable protein levels to SS and MCT, while minimizing reflex tearing. Our method has been tested using tear proteomics, but may also be useful for point of care diagnostics and other applications.

## Materials and Methods

### SCL Protein Quantification

Senofilcon A (Oasys, Acuvue), Nesofilcon A (Biotrue ONEday, Bausch & Lomb), Balafilcon A (Purevision 2, Bausch & Lomb), and Etafilcon A (1-Day Moist, Acuvue) lenses with a spherical equivalent (SE) power between -0.50D and -1.50D were tested in triplicate. Lenses were soaked for 1 h in a simulated human tear protein mixture (HTPM) (human albumin, human lactoferrin, and human lysozyme, 2/2/0.1, w/w/w in Milli-Q H_2_O) to a final concentration of 9 mg/mL. Lenses were removed from the protein solution with forceps and were lightly touched to a Kimwipe to wick away excess fluid. Lenses were then placed in a microcentrifuge tube with 400 μL phosphate buffered saline (PBS, Genesse Scientific), 4M guanidine (GoldBio), or 10% HPLC grade acetone (Fischer Chemical) and sonicated for 10 min to desorb proteins from the lens. Total protein present in the solution was then measured using the Pierce BCA Protein Assay kit (ThermoFisher Scientific) referencing HTPM in 4M guanidine as the standard.

### SCL Total Protein Capture Time Course

Etafilcon A lenses were fully submerged and soaked in 400 μL HTPM for 5 min, 1 h, 4 h, and 16 h. After incubation, excess fluid was wicked using a Kimwipe, protein was removed with 4M guanidine and sonication, and total protein was measured as previously described.

### SCL Total Protein Capture by Dioptric Power

-8.00DS, -0.75DS, and +2.00DS Etafilcon A lenses were removed from their blister packs with forceps and touched to a Kimwipe. Individual lenses were transferred to microcentrifuge tubes and submerged in 400μL HTPM for 1 h. After incubation, excess fluid was wicked using a Kimwipe, then captured protein was removed with 4M guanidine and sonication, and total protein was measured as previously described.

### Human Subject Enrollment & Study Design

Human subjects research was performed in accordance with the Declaration of Helsinki; approval was granted by the Internal Review Board at Brigham Young University (IRB2022-166). Samples were collected at Alpine Vision Center (AVC) (Saratoga Springs, UT). Subjects were educated on the purposes, risks, and benefits of the study. Informed consent was obtained before subject enrollment and privacy rights of human subjects were observed.

Enrollment was based on predetermined inclusion and exclusion criteria. Subjects who were 18 years or younger and pregnant women were excluded from the study. Only subjects with a tear meniscus height >0.3mm were allowed to participate. Three female and three male subjects between the ages of 21 and 29 were recruited. Of these subjects, two males and one female reported having previously used SCLs.

At the beginning of the study, subjects were briefly instructed on SS, MCT, and SCL tear sampling methods. Each subject then had photos taken of their eyes, answered a pre-sampling questionnaire, donated tears by SS, MCT, or SCL, repeated anterior segment photos, answered a post-sampling questionnaire, and waited 45 min before repeating the cycle until all 3 sampling methods were performed. All samples were collected by an optometrist; subjects were not permitted to sample their own tears.

### Bulbar Conjunctival Injection (BCI)

Assessment Photographs of the inferior temporal bulbar conjunctiva were taken using a Keratograph 5M (Oculus) before and after each sampling method. Photos were printed and each subject’s pre- and post-sampling photos were randomly placed side by side. Photos blinded for sampling method were shown to three optometrists who determined which photo in each pair had greater BCI. All photos pairs were scored. Photo pairs with greater BCI before tear sampling were counted as “-1” and those with greater BCI after tear sampling were counted as “1”.

Subjective Tear Sampling Method Assessment Subjects were given a brief overview of all sampling methods at the beginning of the study. Before donating each tear sample, subjects were asked to quantify their anxiety about the tear sampling method on a scale of 0 (no anxiety) to 10 (extreme anxiety). After sampling, the subjects were given a questionnaire assessing their anxiety of repeating the method, discomfort, and difficulty, each on a scale of 0 (none) to 10 (extreme). Subjects were also asked if they would be willing to repeat the sampling method if it could provide useful information about their eye health. If the subject was not willing to repeat the experiment, they were asked to explain. Finally, subjects were asked to rate their overall tear sampling experience from 0 (terrible) to 10 (excellent).

At the conclusion of the study, subjects were asked to preferentially rank order the tear sampling methods from 2 (most preferred) to 0 (least preferred) and explain their answer. Subjects answered questionnaires individually and privately.

### Tear Sampling

#### SS

Wearing nitrile powder-free exam gloves (NIGHT ANGEL, Adenna), the optometrist inserted the tip of a SS (Gulden Ophthalmics) between the lid and the globe of the inferior temporal portion of the subject’s eye. After 5 min, the optometrist donned a new set of gloves, removed the SS, then placed it in a microcentrifuge tube.

#### MCT

Subjects were seated at an SL-D2 slit lamp (Topcon) and instructed to look in superior nasal gaze. A 5μL Microcaps MCT (Drummond) was gently positioned into the inferior temporal tear prism until tears were drawn approximately half-way up the tube. The optometrist attempted to minimize contact with the ocular surface. Tears were then pushed out of the MCT into a microcentrifuge tube using a rubber bulb.

#### SCL

Using new gloves for each lens, the optometrist placed Etafilcon A SCLs (−0.50 DS) on each eye. After 5 min, the optometrist donned a fresh set of gloves, removed the lenses, and placed them in microcentrifuge tubes.

All samples were cold-chain transported on ice and stored at -80 °C. All sampling was done without anesthesia.

### MS Sample Preparation

After thawing all samples, SSs were cut into 2 mm squares with clean scissors. 400 μL 4M guanidine in MS-grade water was then added to each SS, MCT, and SCL sample. All samples were sonicated at room temperature for 10 min, then incubated at 100 °C for 5 min. Total protein was then measured as described previously with standards prepared in 4M guanidine. All tear samples were normalized to 20 μg per sample.

Next, 250 mM tris (2-carboxyethyl) phosphine hydrochloride (TCEP, ThermoFisher Scientific) was added to each sample at volumes sufficient to make a 5 mM final concentration. 250 mM 2-Chloroacetamide (CAA, Acros Organics) was then added to make a final concentration of 15 mM. Samples were then placed on a heat block at 100 °C for 5 min. The entire reaction solution was transferred to a 30 kD Nanosep filter (Pall, Port Washington, NY), centrifuged at 14,000x g for 10 min, and washed with 300 μL 25 mM triethylamine bicarbonate (TEAB, pH 8.5, ThermoFisher Scientific). 100 μL TEAB was mixed with 1 μg/μL trypsin (ThermoFisher Scientific) in the volume above the filter, the samples were pulsed in the centrifuge for 2 seconds, and then placed on a shaker in a 37 °C incubator for 14 h. Samples were then centrifuged at 14,000x g for 30 min before addition of 100 μL 25 mM TEAB pH 8.5 and repeat centrifugation for 30 min. The filtrate was then transferred to vials and labeled using the TMT10-plex Isobaric Label Reagent Set (ThermoFisher Scientific). Two TMT 10-plexes were created, each consisting of 9 individual tear samples and 1 pooled sample.

### Mass Spectrometry

Tear peptides were resuspended in OrbiA (3% ACN, 0.1% FA, 96.9% Optima-LC/MS grade H2O, [all chemicals from Fisher Chemical]) to a concentration of 1 μg/μL. Samples were analyzed with online nanoflow (300 nl/min) liquid chromatography tandem mass spectrometry (LC-MS/MS) using an Ultimate 3000-RSLC nano HPLC system (ThermoFisher Scientific) coupled to an Orbitrap Fusion Lumos MS (ThermoFisher Scientific). 6 μL of each sample was separated with a 75 μm inner diameter (360 μm outer diameter), 25 cm in length microcapillary column packed with 2 μm C18 beads heated to 35 °C and ionized using an electrospray emitter tip (10 μm). HPLC solvent A (OrbiA) and solvent B (OrbiB, 80% ACN, 0.1% FA, 19.9% MS grade H2O). Each sample ran for 266 min with the following gradient at 300 nl/min: 0-2 min, 5% B; 2-231 min, 5 to 32% B; 231-244 min, 32 to 42% B; 244-256 min, 42 to 99% B; 256-266 min, 99% B; the separation gradient was followed with a seesaw wash: 266-269 min, 99 to 2% B; 269-271 min, 2% B; 271-273 min, 2 to 100% B; 273-276 min, 100% B; 276-279 min, 100 to 2% B; 279-281 min, 2 to 100% B; 281-284 min, 100% B; 284-286 min, 100-0% B; 286-288 min, 0% B.

The Orbitrap Fusion Lumos MS was operated in data-dependent mode with a 3 second cycle time to acquire CID MS/MS scans. MS1 data was acquired by orbitrap with resolution of 120,000. The following filters were used to select MS2 scans: precursor range, 400-1400 m/z; monoisotopic peak determination, peptide; intensity threshold, 5.0 × 103; theoretic precursor fit threshold, 70% with a 0.5 m/z fit window; charge states, 2-6; dynamic exclusions, precursor exclusion after 1 time for 60 sec. Selected precursors were activated with a fixed 35% collision energy, and MS2 data was detected with the ion trap at a scan rate of 125,000 Da/sec set to 50 msec max injection time. CID collision energy was set to 30% with a 10 ms CID activation time. As part of the TMT 10-plex workflow, precursors ranging from 400-1600 m/z with an exclusion window of 25 ppm were selected for MS3 fragmentation. Synchronous precursor selection was set to 10, and precursors were activated with 65% (normalized) HCD. Orbitrap scans with a range of 110-500 were collected at a resolution 50,000 with a max injection time of 105 msec.

### Data Analysis

Statistical analysis of in vitro SCL testing, subject questionnaire data, and inter-method BCI comparisons were performed using one-way ANOVA. The absolute value of the average BCI score was used to calculate statistical significance between sampling methods. One-sample t-tests were used to calculate significance in scored pre- and post-sampling BCI for each individual method.

MS data was analyzed using Peaks software (Bioinformatics Solutions Inc.). Spectrum filter settings were set to false discovery rate (FDR) of 1% (−10logP ≥ 26.4) and quality ≥ 8.7. Protein filter settings were significance ≥ 0, fold change ≥ 1, and at least 1 unique peptide. All spectra with intensity < 1 × 102 were excluded and only proteins present in ≥ 3 subjects per sampling method were counted. For quantitative analysis, normalization of each TMT 10-plex was performed separately by grouping sampling methods and normalizing to the average signal intensity of the pooled sample. All data was log2 transformed and protein fold change was calculated for each subject. Sampling methods were then compared by performing one-sample, paired, two-tailed t-tests for all proteins. FDR was accounted for by adjusting p-values with the Benjamini-Hochberg equation (FDR = 0.25) with adjusted p-values <0.05 considered significant. Groups of proteins unique to each sampling method and shared between methods were analyzed separately for functional enrichment relative to the combined list of all identified proteins using the STRING database [12].

The raw data is available on the Mendeley data repository [13].

## Results

To develop a method for SCL tear sampling, we began by selecting SCL candidates. Senofilcon A, Nesofilcon A, Balafilcon A, and Etafilcon A were chosen to represent a broad spectrum of chemical compositions and characteristics (Fig. S1).

We first compared the ability of the selected SCLs to capture human tear proteins in vitro. Each SCL (n=3) was exposed to a simulated human tear protein mixture (HTPM) that contained human albumin, lactoferrin, and lysozyme at a physiological total protein concentration [1]. After protein capture, three protein removal methods were tested: a saline solution (1x PBS), a chaotropic agent (4M guanidine), and an organic solvent (10% acetone) (Fig. 1A). The etafilcon A SCL and guanidine combination provided the highest protein yield, with a ∼3-fold increase in recovered protein compared to other SCL/chemical combinations.

**Fig. 1.**
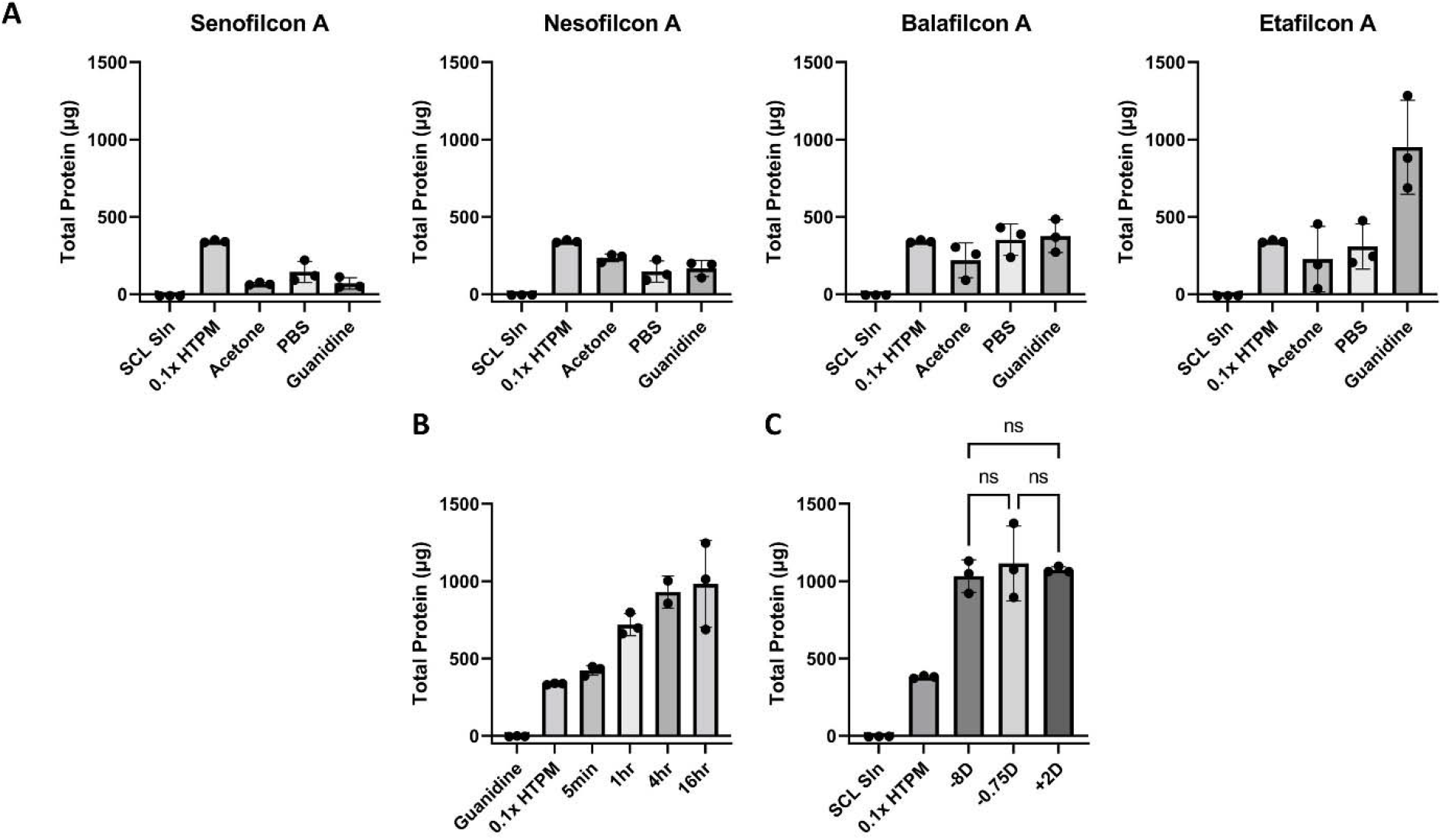
SCL protein sampling in vitro. A) Total protein of Nesofilcon A, Balafilcon A, Etafilcon A, and Senofilcon A lenses incubated with PBS, 10% acetone, and 4M guanidine, B) Total protein from Etafilcon A lenses soaked in HTPM for 5 min, 1 hr, 4 hrs, and 16 hrs, C) Total protein by SCL power (−0.75DS, -8.00DS, and +2.00DS) after a 1 hr incubation in HTPM. Error bars represent standard deviation, n=3 for all experiments. NS = not significant

To estimate how long SCLs should be in contact with tears on the eye, we assessed the impact of SCL protein capture time in vitro by soaking SCLs in HTPM for 5 min, 1 h, 4 h, and 16 h, followed by a 10-min incubation in 4M guanidine (Fig. 1B). A 5-min SCL soak in HTPM captured an average of 450±37 μg of protein. Protein capture increased with time and yielded an average of 1.07±0.28 mg at 16 hours. However, since several hundred μg is ample for most MS proteomics experiments, we concluded that a 5 minute on eye sampling was sufficient. Notably, this sample time allows for feasible tear film protein collection during routine eye exam visits.

Given that SCL dioptric power is related to lens thickness, and thicker lenses could potentially absorb more protein, we assessed whether SCL dioptric power affected total protein capture. As shown in Fig. 1C, differences in dioptric power showed no statistically significant difference in collected protein yield. We conclude that yields of captured protein are largely insensitive to SCL dioptric power.

After analyzing the data from our in vitro experiments, we created a SCL protein sampling method that would be used to analyze human tear film. Etafilcon A lenses are placed on the subject’s eye for 5 minutes. The lens is removed, placed in 400μL 4M guanidine, and sonicated for 10 minutes. This solution is then ready for downstream analysis. We analyzed the SCL elution using MS as described in Materials and Methods (Fig. 2A).

**Fig. 2.**
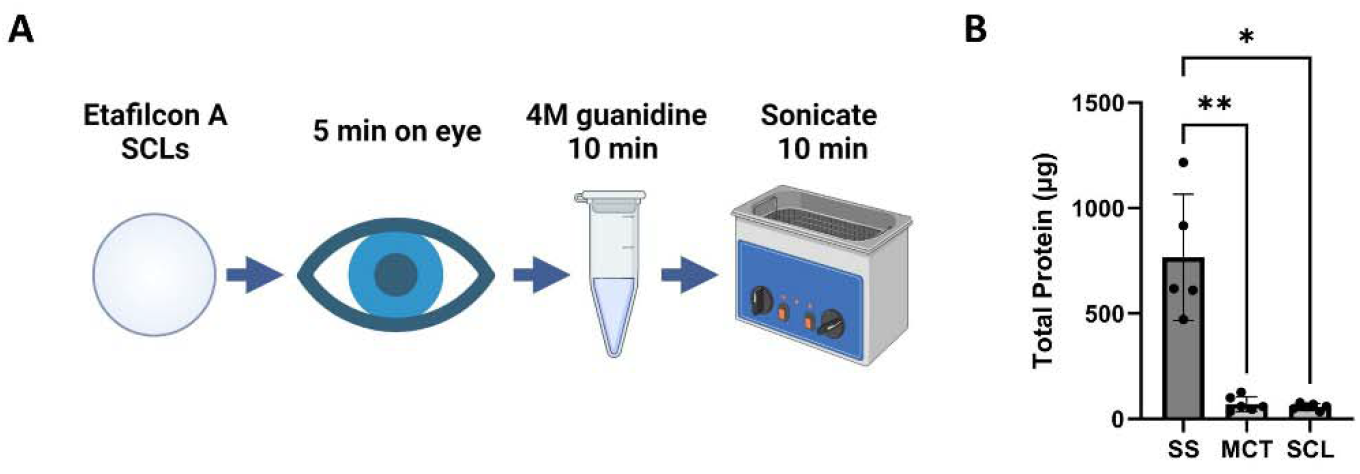
SCL human tear sampling. A) Method overview, B) Total protein from human subjects for 1 eye using SS, MCT, and SCL sampling. Error bars represent standard deviation, n=6. * = p-value ≤0.05, ** = p-value ≤ 0.01

We tested our SCL tear film sampling method on 6 human subjects: 3 male and 3 female subjects between the ages of 21 and 29. Each subject donated tears by SS, MCT, and SCL, yielding averages of 260±60, 40±28, and 60±31 μg total tear protein, respectively (Fig. 2B). As expected, SS yielded the largest amount of protein. The SCL and MCT methods gave similar results. We attribute the difference in total protein between SS and MCT/SCL mostly to sampling volume, since SS methods have a significantly larger tear sampling capacity. For SCL sampling, differences between in vitro and in vivo sampling may reflect differential protein capture in HTPM versus individual subject tear proteins, or partial mechanical protein removal due to wiping of the SCLs by the eyelids.

One potential source for a greater sample volume in SS tear sampling could be ocular irritation. Importantly, eye irritation from tear sampling can alter the proteomic profile and increase subject reluctance for future studies [14,15]. Hence, we assessed conjunctivitis, a known sign of ocular irritation [16]. Objective signs of conjunctivitis were determined for each subject by comparing bulbar conjunctival injection (BCI) before and after each tear film sampling method. Three optometrists performed a blinded, pairwise comparison of BCI from pre- and post-sampling photos. As shown in Fig. 3B, ocular surface irritation was significantly higher post-SS sampling compared to MCT or SCL. We conclude that differences in pre- and post-sampling BCI for SS are significant and demonstrate irritation from sampling, while differences for MCTs or SCLs are indistinguishable and thus do not induce ocular irritation, as observed by BCI (Fig. 3C).

**Fig. 3.**
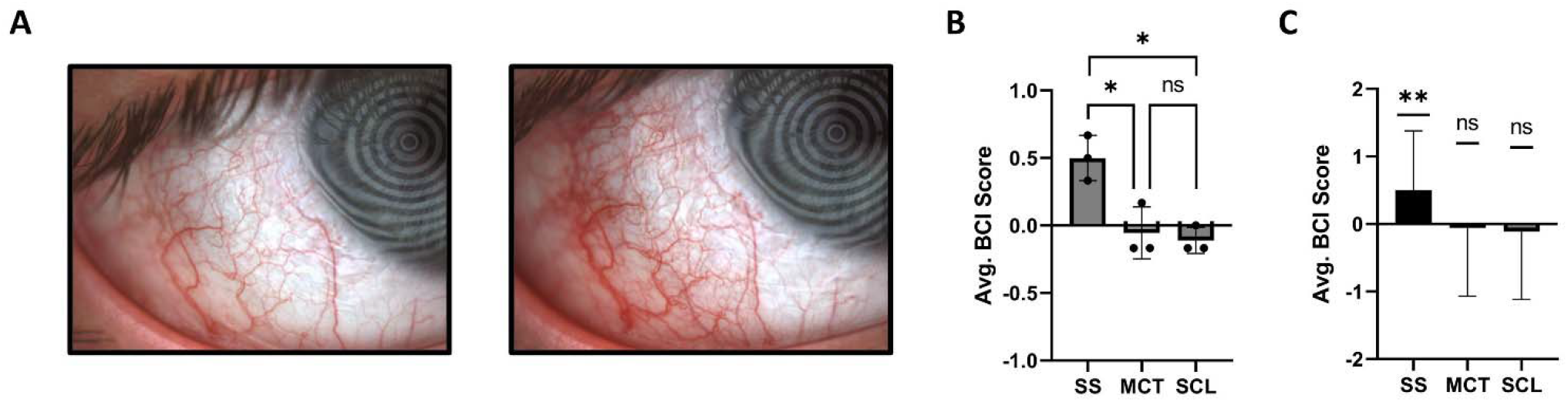
Objective bulbar conjunctival injection (BCI). A) Photos demonstrating pre-(left) and post-(right) SS sampling, B) Scoring of all pre- and post-sampling photos for all subjects and methods by three optometrists in a blinded pairwise comparison. BCI Score: Photo pairs scored with pre-sampling photos having greater BCI were counted as “-1”. Post-sampling photos scored with greater BCI were counted as “1”, C) Average BCI score for individual sampling methods analyzed by one-sample t-tests. Error bars represent standard deviation, n=12. * = p-value ≤0.05, ** = p-value ≤ 0.01

Subject cooperation is required for effective sampling. Hence, we sought to understand the subject’s perspective on each sampling method. In our small study, subject questionnaire responses revealed anxiety about all methods (Fig. 4A). Post sampling, subjects viewed MCT and SCL with less anxiety than SS (Fig. 4B). On average, subjects reported the lowest level of discomfort with MCT (Fig. 4C). Subject assessment of tear sampling difficulty was also queried. Responses were varied; there was no statistically significant difference between methods (Fig. 4D). Finally, subjects were asked to numerically rate their overall experience with each method. Subjects reported the best experience with MCT and the worst experience with SS (Fig. 4E).

**Fig. 4.**
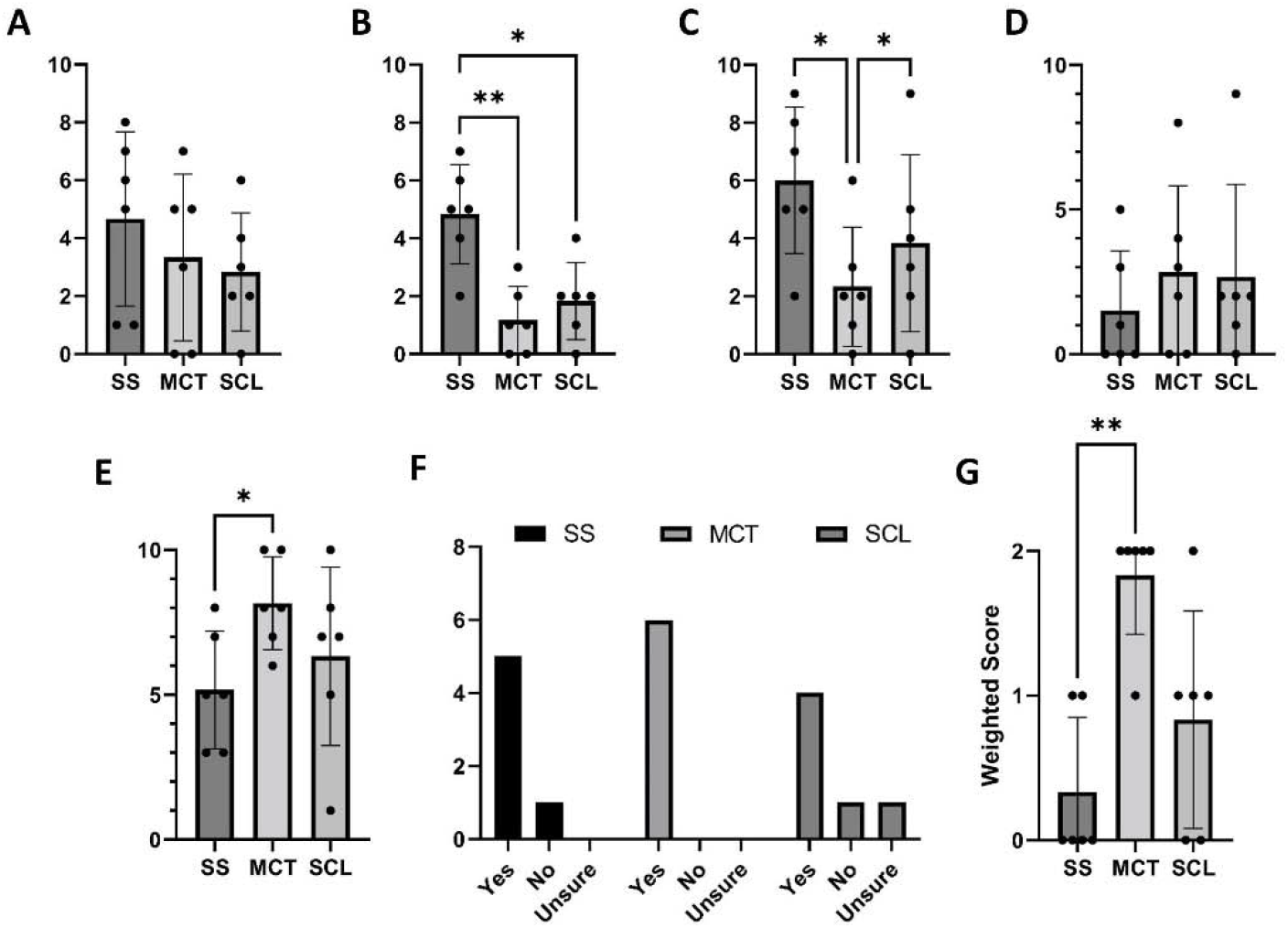
Subject tear sampling questionnaire responses. A) Anxiety pre-sampling, B) Anxiety post-sampling, C) Discomfort during sampling, D) Difficulty of sampling, E) Willingness to repeat the sampling method if it could provide useful health information, F) Overall experience, G) Rank order of preferred tear sampling method. All questionnaires were graded on a scale of 0 = none, to 10 = extreme, except for willingness to repeat (graded “yes”, “no”, or “unsure”), and overall experience (graded 0 = terrible, to 10 = excellent). Error bars represent standard deviation, n=6. * = p-value ≤0.05, ** = p-value ≤0.01

Tear film biomarker discovery or diagnostic screening would require subject/patient acceptance of tear sample collection. When asked if they would be willing to repeat each tear sampling method if it could provide useful eye health information, most subjects responded positively. One subject reported that they would refuse SS due to discomfort, 1 subject (who had not previously worn contact lenses) reported that they would refuse SCL due to perceived sampling difficulty, and 1 subject who reported “unsure” further explained that they would prefer the SCL method if they could perform SCL insertion and removal themselves (Fig. 4F). Overall, most subjects reported the best experience with MCT and were willing to repeat any method if it could provide useful ocular health information.

At the conclusion of the subject study, subjects were asked to compare tear sampling methods and rank them by preference. Subjects ranked MCT as most preferred, followed by SCL, with SS as the least preferred method (Fig. 4G). Subjects further commented that they perceived MCT as the fastest, least invasive, most comfortable, and easiest method. Two of three subjects who were SCL wearers commented that they would choose SCL over MCT if they could insert and remove the SCLs themselves.

Finally, we assessed the identifiable proteins present in tear samples for each sampling method. Using MS proteomics, we identified 482, 448, and 387 total proteins out of the SS, MCT, and SCL subject samples, respectively (Fig. 5, Table S1). The majority (386 proteins) were shared between all methods. Furthermore, 36 proteins were unique to SSs, while MCTs and SCLs had only 2 and 1 unique proteins, respectively. Additionally, 60 proteins were shared between SS and MCT with no other shared proteins between sampling methods. Functional enrichment analysis of unique and shared proteins between all methods were analyzed using the STRING database. Our queries did not return any statistically significant results for groups of unique or shared protein species found across the different tear sampling methods.

**Fig. 5.**
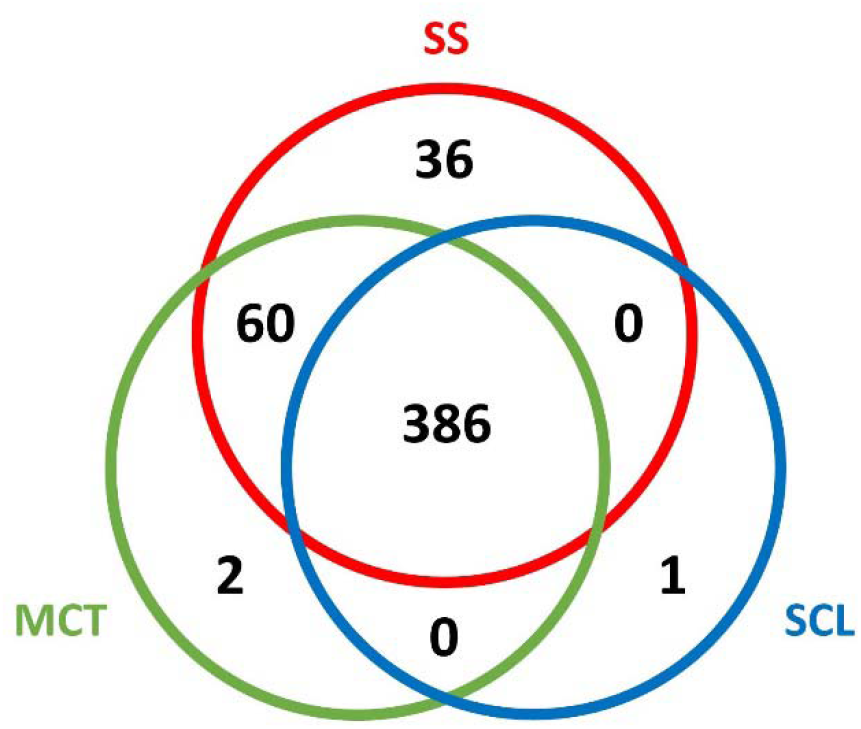
Total protein identifications by MS for tear sampling methods. SS = red, MCT = green, SCL = blue

We assessed identified proteins in all tear sampling methods for previously reported reflex tear markers [15]. Our proteomics data showed zymogen granule protein 16 homolog B (ZG16B) decreased significantly in SCL sampled tears compared to SS and MCT for all subjects (Table 1), suggesting that SCL sampling elicits less reflex tearing. Comparing SS to MCT revealed significant levels of different reflex tear proteins for both methods. Surprisingly, polymeric immunoglobulin receptor (PIGR) was elevated in SCLs relative to MCT only for the SCL non-wearer subgroup (Table S2). As expected, Lysozyme C (LYSC) was elevated in SCL sampling since SCLs preferentially adsorb LYSC [17]. However, lactoferrin (TRFL) and lipocalin-1 (LCN1) were significantly reduced with SCL sampling. Since TRFL accounts for over 25% of total tear proteins [18] and LCN1 is the second most concentrated tear protein [19], a reduction in TRFL and LCN1 should reduce protein abundance suppression in MS; a potential benefit of SCL sampling [20,21]. Thus, SCL sampling shows fewer signs of reflex tearing and could aid in the discovery of low abundant biomarkers by reducing levels of highly abundant proteins. Future studies with larger sample sizes will be needed to elucidate proteomic differences in tear sampling methods.

**Table 1.** Quantitative changes in reflex selected tear proteins between sampling methods

| Method Comparison | Reflex Tear Change [15] | Accession\|Protein <sup>a</sup> | Avg. FC <sup>b</sup> | Adj. p-value <sup>c</sup> |
| --- | --- | --- | --- | --- |
| SS/MCT | Increase | P02768\|ALBU | 2.89 | NS |
|  | Increase | Q16378\|PROL4 | 0.37 | 0.019 |
|  | Increase | Q96DA0\|ZG16B | 0.65 | NS |
|  | Decrease | P10909\|CLUS | 0.30 | 0.013 |
|  | Decrease | P01037\|CYTN | 0.34 | 0.014 |
|  | Decrease | P01876\|IGHA1 | 0.47 | 0.049 |
|  | Decrease | P01833\|PIGR | 0.27 | 0.012 |
|  | Decrease | O75556\|SG2A1 | 0.35 | 0.016 |
|  | NS | P31025\|LCN1 | 1.13 | NS |
|  | NS | P61626\|LYSC | 0.66 | NS |
|  | NS | P02788\|TRFL | 0.56 | 0.015 |
| SS/SCL | Increase | P02768\|ALBU | 4.40 | NS |
|  | Increase | Q16378\|PROL4 | 1.53 | NS |
|  | Increase | Q96DA0\|ZG16B | 3.26 | 0.005 |
|  | Decrease | P10909\|CLUS | 2.37 | NS |
|  | Decrease | P01037\|CYTN | 2.55 | NS |
|  | Decrease | P01876\|IGHA1 | 3.12 | NS |
|  | Decrease | P01833\|PIGR | 2.80 | NS |
|  | Decrease | O75556\|SG2A1 | 3.40 | 0.032 |
|  | NS | P31025\|LCN1 | 3.93 | 0.003 |
|  | NS | P61626\|LYSC | -1.57 | 0.002 |
|  | NS | P02788\|TRFL | 3.20 | 0.008 |
| MCT/SCL | Increase | P02768\|ALBU | 1.51 | NS |
|  | Increase | Q16378\|PROL4 | 1.17 | NS |
|  | Increase | Q96DA0\|ZG16B | 2.61 | 0.005 |
|  | Decrease | P10909\|CLUS | 2.07 | NS |
|  | Decrease | P01037\|CYTN | 2.21 | NS |
|  | Decrease | P01876\|IGHA1 | 2.65 | NS |
|  | Decrease | P01833\|PIGR | 2.54 | 0.032 |
|  | Decrease | O75556\|SG2A1 | 3.05 | 0.025 |
|  | NS | P31025\|LCN1 | 2.80 | 0.016 |
|  | NS | P61626\|LYSC | -2.23 | 0.001 |
|  | NS | P02788\|TRFL | 2.64 | 0.014 |

<sup>a</sup> Albumin (ALBU), proline-rich protein 4 (PROL4), zymogen granule protein 16 homolog B (ZG16B), clusterin (CLUS), cystatin-SN (CYTN), immunoglobulin heavy constant alpha 1 (IGHA1), polymeric immunoglobulin receptor (PIGR), mammaglobin-B (SG2A1), lipocalin-1 (LCN1), lysozyme C (LYSC), and lactotransferrin (TRFL)
<sup>b</sup> FC = Fold Change
<sup>c</sup> Statistical significance was tested using the Benjamini-Hochberg equation. NS = not significant

## Discussion

All tear sampling methods involve foreign bodies touching the ocular surface, and foreign bodies often cause irritation, risk of injury, and the potential confounding effects of reflex tearing [22,23]. Thus, optimal tear sampling involves a soft and comfortable foreign body to reduce negative side effects. In particular, SS are known to cause ocular surface irritation and subsequent reflex tearing [24]. In fact, SS are so irritating that topical anesthesia is commonly used with SS in dry eye testing to reduce reflex epiphora [25]. However, anesthesia is a confounding variable and not always an appropriate solution to eye irritation.

Not surprisingly, SCLs show significantly less ocular surface irritation than SS since SCLs are designed for comfort and safety on the eye. Similar to SCLs, our BCI data shows MCT irritate the eye less than SS. However, it is important to note that for MCT and SCL sampling, irritation, reflex tearing, and subject preference are dependent upon the skill of the person carrying out the sample collection rather than the method itself. Unlike SCLs, MCT sampling takes significant dexterity to perform without irritating the ocular surface and should be performed by a specialist with access to a slit lamp [24]. In addition, the person sampling tears using SCLs only needs experience in SCL insertion and removal; subjects can sample their own tears.

In our study, subjects reported that MCT was the most preferred tear sampling method. However, this data conflicts with previous reports, including a recent study in which MCT was ranked the least comfortable of four different methods, including SS [26,27]. A possible explanation for this discrepancy was that in our study, an optometrist trained in MCT sampling collected tears using a slit lamp. Not all researchers use slit lamp microscopy for MCT sampling, which can affect the subject’s experience and increase risk of injury and discomfort. It is also important to note that MCT samples are more difficult to collect from low tear volume dry eye disease (DED) subjects, a condition which increases linearly with age [28]. In this study, all subjects were young and had a healthy tear volume prior to sampling [29]. Hence, while subjects here responded favorably to MCT sampling, an older subject population, sampling without a slit lamp, and/or less practiced hands carrying out the sample collection could yield a different result.

Each tear sampling method may have advantages over other methods, depending on the subject demographic and study design. For example, SCLs may be preferred for researchers or subjects with previous SCL experience. To our knowledge, SCL sampling is also the only known method where both researcher sampling and subject self-sampling are reasonable options. Importantly, the option for self-sampling creates the potential for at-home tear sampling and would allow for subject to insert the SCLs in the morning prior to their eye exam as a method to collect more protein throughout the day. This a strategy could make SCLs the tear sampling method of choice for low abundance biomarkers.

To analyze proteins collected by the different sampling methods, we used MS proteomics, the method typically applied for tear biomarker identification [9]. Our proteomic analysis confirmed that all tear sampling methods capture a common set of proteins representing the majority of all proteins sampled by each method. Consistent with previous reports, SS sampling returned higher protein identifications compared to MCT [8,9]. To rule out the possibility that this difference simply reflects the observed larger sample volumes and thus higher total protein, our MS analysis normalized for total protein concentration between methods. Notably, even when corrected for differences in protein yield, the MS data still showed a significant subset of proteins unique to SS. Previous studies comparing SS and MCT proteomics show similar subsets unique to SS, including a label-free experiment reported by Nättinen et al. where 850 proteins were identified, 80 were unique to SS, 9 were unique to MCT, and 761 were shared [8,30]. We note that in their study, total sampled protein was not normalized between methods (Avg MCT: 19.7 μg, Avg SS: 199 μg).

Since the reflex tear profile is reportedly different than that of basal tears [14,15,24], we evaluated the respective tear proteomes for proteins known to be associated with reflex tearing. As there is currently no reference proteome for reflex tears, we relied mainly on the recent report by Perumal, et. al [15]. It is important to note that in their study both basal and reflex tears were collected using MCTs and reflex tearing was stimulated using onion vapors. As reflex tearing has many different triggers [31], proteomic differences may exist depending on how reflex tearing is stimulated. Thus, more research is needed to assess reflex tear stimulation subtypes and validate their associated proteomes.

In our hands, ZG16B is decreased in SCL sampled tears compared to both SS and MCT. Although the role of secretory lectin protein ZG16B in tears remains largely unknown, it has been proposed that its expression in reflex tears is connected to neuronal stimulation of the lacrimal gland [32]. While previous reports state that SS sample reflex tears and MCT sample basal tears [24,33], these data indicate that SCLs may improve basal tear sampling by avoiding reflex tear stimulation compared to SS and MCT. One potentially contrary finding is the measured abundance of PIGR, a putative reflex tear protein which could suggest ocular irritation below our detectable BCI threshold. However, PIGR was only significant in SCL non-wearers, and subjects who have not previously worn SCLs are expected to show greater signs of irritation relative to established SCL wearers. Overall, our combined clinical and proteomic data supports that SCL tear sampling shows the fewest signs of reflex tearing and is an improved method for sampling basal tears, particularly in SCL wearers.

We note that our ocular BCI findings support previous reports that SS is irritating to the ocular surface [7] and stimulates reflex tearing [24,34]. Our data suggests that unique SS proteins may be linked to irritation and ocular surface damage, a hypothesis that has been made previously [35]. However, our studies also show a set of 60 proteins shared by SS and MCT that were not detected by SCL sampling. While it is possible that SCLs simply did not capture this subset of proteins, our ZG16B findings suggest that biochemical processes associated with reflex tearing might be stimulated with MCT sampling, even without visible signs of irritation. Taken together, these findings support SCL sampling as a preferred method for basal tear sampling.

This study has several important limitations. First, our pilot study had a small sample size. Second, the researcher sampling tears had practiced MCT sampling in preparation for this study, which may have affected both proteomic and subject questionnaire results. Third, all subjects were under 30 years of age. Since tear volume decreases linearly with age, older subjects should be included in sampling assessments in future larger studies.

SCL tear sampling is an accessible method for analysis of tear film proteins. Our study highlights the need for careful selection of tear sampling method. The desired tear type, effects on the subject, and available resources in both personnel and equipment are all important considerations. SS are easy to use, but irritate the eye and induce reflex tearing. When performed by a dexterous researcher with access to a slit lamp, subject cooperation with MCT can be high. However, MCT may be inappropriate for DED subjects. SCLs are an easily implemented tear sampling method appropriate for DED subjects that do not require additional equipment. More importantly, SCL sampling shows low BCI relative to SS and the lowest levels of reflex tear protein ZG16B. We conclude that SCLs provide an accessible method for tear sampling with minimal accompanying ocular irritation.

## Data Availability

All data produced are available online on the Mendeley data repository.

https://data.mendeley.com/datasets/f4vggkmcdn/draft?a=82975819-b7d7-486a-9db2-76919a7a7cfb

## Acknowledgements

We would like to thank Dr. Steven Weaver, Dr. Carlan Reese, and Dr. Jared Freestone at AVC.

## Declarations

### Ethics approval

Approval for research involving human subjects was granted by the Internal Review Board at Brigham Young University (IRB2022-166).

