## Supplementary material for "Human Tear Film Protein Sampling Using Soft Contact Lenses": Fig. S1

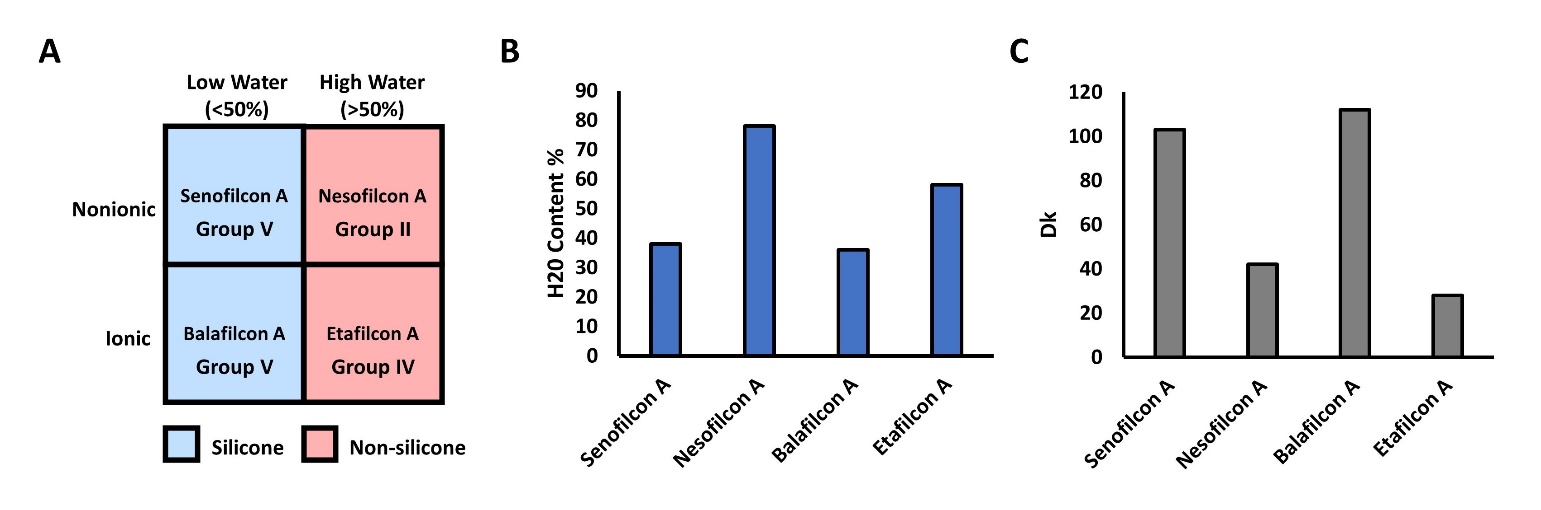

Specifications of SCLs used. A) Hydrogel polymers by FDA grouping, B) Water content by polymer, C) Oxygen transmissibility (Dk) by polymer. Data is shown as reported by contact lens manufacturers
