## Supplementary material for "Human Tear Film Protein Sampling Using Soft Contact Lenses": Table S2

**Table S2. Quantitative changes in reflex selected tear proteins between sampling methods for SCL wearers and non-wearers**

|  |  |  | SCL Non-wearers | | SCL Wearers | |
| --- | --- | --- | --- | --- | --- | --- |
| Method Comparison | Reflex Tear Change [15] | Accession\|Protein ^a^ | Avg. FC ^b^ | Adj. p-value ^c^ | Avg. FC ^b^ | Adj. p-value ^c^ |
| SS/MCT | Increase | P02768\|ALBU | 4.16 | 0.018 | 1.61 | NS |
|  | Increase | Q16378\|PROL4 | 0.31 | NS | 0.43 | 0.033 |
|  | Increase | Q96DA0\|ZG16B | 0.30 | 0.024 | 0.99 | NS |
|  | Decrease | P10909\|CLUS | 0.48 | NS | 0.11 | 0.010 |
|  | Decrease | P01037\|CYTN | 0.48 | NS | 0.20 | 0.017 |
|  | Decrease | P01876\|IGHA1 | 0.39 | NS | 0.55 | NS |
|  | Decrease | P01833\|PIGR | 0.20 | NS | 0.33 | 0.023 |
|  | Decrease | O75556\|SG2A1 | 0.39 | NS | 0.31 | 0.031 |
|  | NS | P31025\|LCN1 | 1.02 | NS | 1.24 | NS |
|  | NS | P61626\|LYSC | 0.55 | NS | 0.76 | NS |
|  | NS | P02788\|TRFL | 0.53 | NS | 0.59 | 0.030 |
| SS/SCL | Increase | P02768\|ALBU | 5.57 | NS | 3.23 | 0.040 |
|  | Increase | Q16378\|PROL4 | 1.23 | NS | 1.84 | 0.015 |
|  | Increase | Q96DA0\|ZG16B | 2.99 | NS | 3.52 | 0.003 |
|  | Decrease | P10909\|CLUS | 2.59 | NS | 2.15 | NS |
|  | Decrease | P01037\|CYTN | 2.73 | NS | 2.37 | NS |
|  | Decrease | P01876\|IGHA1 | 3.93 | NS | 2.30 | NS |
|  | Decrease | P01833\|PIGR | 3.33 | NS | 2.27 | NS |
|  | Decrease | O75556\|SG2A1 | 3.37 | NS | 3.43 | 0.002 |
|  | NS | P31025\|LCN1 | 3.66 | NS | 4.19 | 0.005 |
|  | NS | P61626\|LYSC | -1.87 | 0.016 | -1.28 | 0.008 |
|  | NS | P02788\|TRFL | 2.83 | 0.041 | 3.56 | 0.010 |
| MCT/SCL | Increase | P02768\|ALBU | 1.40 | NS | 1.62 | NS |
|  | Increase | Q16378\|PROL4 | 0.92 | NS | 1.41 | NS |
|  | Increase | Q96DA0\|ZG16B | 2.69 | 0.021 | 2.53 | 0.025 |
|  | Decrease | P10909\|CLUS | 2.11 | NS | 2.03 | NS |
|  | Decrease | P01037\|CYTN | 2.25 | NS | 2.17 | NS |
|  | Decrease | P01876\|IGHA1 | 3.54 | NS | 1.75 | NS |
|  | Decrease | P01833\|PIGR | 3.13 | 0.028 | 1.94 | NS |
|  | Decrease | O75556\|SG2A1 | 2.98 | NS | 3.12 | 0.006 |
|  | NS | P31025\|LCN1 | 2.65 | NS | 2.95 | 0.010 |
|  | NS | P61626\|LYSC | -2.42 | 0.002 | -2.04 | 0.003 |
|  | NS | P02788\|TRFL | 2.31 | 0.035 | 2.97 | 0.015 |

^a^ Albumin (ALBU), proline-rich protein 4 (PROL4), zymogen granule protein 16 homolog B (ZG16B), clusterin (CLUS), cystatin-SN (CYTN), immunoglobulin heavy constant alpha 1 (IGHA1), polymeric immunoglobulin receptor (PIGR), mammaglobin-B (SG2A1), lipocalin-1 (LCN1), lysozyme C (LYSC), and lactotransferrin (TRFL).

^b^ FC = Fold Change

^c^ Statistical significance was tested using the Benjamini-Hochberg equation. NS = not significant
